# Intrafamilial Exposure to SARS-CoV-2 Induces Cellular Immune Response without Seroconversion

**DOI:** 10.1101/2020.06.21.20132449

**Authors:** Floriane Gallais, Aurélie Velay, Marie-Josée Wendling, Charlotte Nazon, Marialuisa Partisani, Jean Sibilia, Sophie Candon, Samira Fafi-Kremer

## Abstract

**Background:** In the background of the current COVID-19 pandemic, serological tests are being used to assess past infection and immunity against SARS-CoV-2. This knowledge is paramount to determine the transmission dynamics of SARS-CoV-2 through the post pandemic period. Several individuals belonging to households with an index COVID-19 patient, reported symptoms of COVID-19 but discrepant serology results.

**Methods:** Here we investigated the humoral and cellular immune responses against SARS-CoV-2 in seven families, including nine index patients and eight contacts, who had evidence of serological discordances within the households. Ten unexposed healthy donors were enrolled as controls.

**Results:** All index patients recovered from a mild COVID-19. They all developed anti-SARS-CoV-2 antibodies and a significant T cell response detectable up to 69 days after symptom onset. Six of the eight contacts reported COVID-19 symptoms within 1 to 7 days after the index patients but all were SARS-CoV-2 seronegative. Six out of eight contacts developed a SARS-CoV-2-specific T cell response against structural and/or accessory proteins that lasts up to 80 days post symptom onset suggesting a past SARS-CoV-2 infection.

**Conclusion:** Exposure to SARS-CoV-2 can induce virus-specific T cell responses without seroconversion. T cell responses may be more sensitive indicators of SARS-Co-V-2 exposure than antibodies. Our results indicate that epidemiological data relying only on the detection of SARS-CoV-2 antibodies may lead to a substantial underestimation of prior exposure to the virus.

## Introduction

Corona Virus Disease 2019 (COVID-19) is a pandemic infection that raises a major concern all around the world[1]. To contain the spread of the virus, several countries have imposed population lockdown[2]. In France, the first cases of COVID-19 were recorded at the end of January 2020[3]. Due to the rapid increase of new cases and mortality, the lockdown was imposed from March 17 to May 11, 2020. Since the lifting of the lockdown, the number of new cases of SARS-CoV-2 has decreased significantly. However, it is not excluded that a second pandemic wave may occur in the future [4].

Estimation of immunizing infections is crucial in helping to predict the post pandemic dynamics of the virus[4]. Serological tests for SARS-CoV-2 have been developed to determine the extent of immunity to the virus[4], and immunity certifications based on these tests have been considered by some European countries and by the US government [5]. Since their availability in France, anti-SARS-CoV-2 antibody assays met a huge demand from people to learn if they are protected. Surprisingly, several individuals belonging to households with an index COVID-19 patient reported symptoms of COVID-19 but remained seronegative although no quarantine measures were respected by the index patient. The absence of antiviral antibodies after exposure has been previously reported for other viral infections. In these cases, the presence of viral specific-T cell responses provided proofs of viral transmission[6-8].

In this study, we investigated the humoral and cellular response against SARS-CoV-2 in seven families who reported serological discordances in their household.

## Methods

### Study subjects

Seven households were enrolled in the study. Each involves at least one index patient with a documented proof of positive reverse-transcriptase polymerase chain reaction (RT-PCR) and /or serological testing for SARS-CoV-2, and at least one contact with a negative SARS-CoV-2 serology. All index patients recovered from a mild form of COVID-19 that occurred between March 2 and March 25, 2020. Clinical history was recorded for index patients and contacts. Blood samples for the present study were collected between May 7 and May 28, 2020. Ten healthy donors, who have not been exposed to COVID-19 patients and who have been tested negative for anti-SARS-CoV-2 antibodies, were enrolled as controls. All the participants gave written informed consent for research according to protocols approved by the institutional review board of Strasbourg University Hospitals (CE-2020-34).

### SARS-CoV-2 RT-PCR and Serological tests

In house real-time reverse transcriptase PCR (RT-PCR) tests for SARS-CoV2 nucleic acid were performed on nasopharyngeal swabs. Primer and probe sequences target two regions on the RdRp gene and are specific to SARS-CoV2. Assay sensitivity is around 10 copies / reaction (Institut Pasteur, Paris, France). Three serological assays were used to determine the presence of anti-SARS-CoV-2 antibodies in index patients and contacts: i) The Abbott Architect SARS-CoV-2 IgG is a chemiluminescent microparticle immunoassay for detection of IgG against the SARS-CoV-2 nucleoprotein; and ii) The Euroimmun Anti-SARS-CoV-2 Assay is an enzyme-linked immunosorbent assay (ELISA) for the detection of IgG against the SARS-CoV-2 S1 domain of the spike protein including the immunologically relevant receptor binding domain (RBD). The two assays were approved by the FDA and the French National Agency of Medicine and Health Products Safety (ANSM) regarding their excellent analytical performances (Abbott Architect assay: sensitivity 100% and specificity 100% [9], and Euroimmun assay: sensitivity 100% and specificity 97.7%)[10]. iii) The Biosynex is a lateral flow assay for detection of IgM and IgG against the SARS-CoV-2 RBD of the Spike protein S. This assay was also approved by the ANSM with excellent analytical performances (specificity 99.4% and sensitivity 95.6% [11]). All of the three assays were tested according to manufacturer’s instructions.

### IFN-1 Enzyme-linked Immunospot assay

T cell immune response against SARS-CoV-2 was investigated by performing an interferon-gamma (IFN-L) enzyme linked immunospot (ELISpot) assay in duplicate on fresh peripheral blood mononuclear cells (PBMCs) isolated from heparin-anticoagulated blood. PBMCs were seeded at 200,000 CD3 positive cells/well and stimulated for 20 +/- 4 hours with overlapping 15-mer peptide pools spanning the sequences of the entire SARS-CoV-2 spike glycoprotein (pools S1 and S2), the nucleoprotein (N), the membrane protein (M), the envelope small membrane protein (E) and the accessory proteins 3A, 7A, 8 and 9B (PepMix™, JPT Peptide Technologies, Strassberg, Germany). To investigate the possibility of pre-existing cross-reactive coronavirus-specific T cells, PBMCs were stimulated in parallel with peptide pools spanning the spike glycoprotein sequences of HCoV-229E (ES1 and ES2) and HCoV-OC43 (OS1 and OS2), respectively. Phytohemagglutinin (PHA) was used as a positive control and culture medium in quadruplicate as a negative control. After colorimetric revelation of IFNγ capture (UCytech, Utrecht, The Netherlands), spots were counted using an ELISPOT reader (AID, Strassberg, Germany). For each condition, the mean number of spot forming cells per million CD3 positive cells was calculated from duplicates after subtraction of the background value obtained from negative controls to determine the frequency of antigen specific T cells. The threshold defining a significant SARS-CoV-2-specific response was set at exceeding 3 standard deviations of the negative control background.

## Results

### Index patients and contacts characteristics

Nine index patients (P1, P2, P3A, P3B, P3C, P4, P5, P6 and P7) recovered from a mild COVID-19 with positive serological testing for SARS-CoV-2 and eight contacts (C1, C2, C3, C4A, C4B, C5, C6 and C7) with negative serological testing for SARS-CoV-2 were enrolled in the study. For each household, blood samples were collected the same day, except for the household 5 (Table 1). The median age of index patients was 45 years (range, 34-65 years) and four (50%) were male. They all had a normal lymphocyte count (table 1). SARS-CoV-2 RT-PCR test was performed for 4 households (4, 5, 6 and 7) in nasopharyngeal specimens. RT-PCR was positive for all the index patients and negative for all the contacts (Table 1). Of the nine index patients, five were healthcare workers and two of them reported a history of contacts with COVID-19 positive patients during their work. Patients reported history of fever (n=5), cough (n=6), headache (n=6), anosmia (n=7), ageusia (n=6), and less often dyspnea (n=2) and myalgia (n=2). The duration of symptoms varied from 2 to 15 days (median 7 days). All families rigorously washed their hands and all except one family (Household 2) avoided hugs and kisses with household members. Only two index patients (P4 and P6) quarantined themselves by dieting separately and/or wearing a mask one and three days after symptom onset, respectively. Of the eight contacts, two (C3 and C6) had no symptoms, four had fever, three had cough, four had headache and one had ageusia. The duration of symptoms varied from 1 to 11 days (median 7 days) (Table 1).

**Table 1.**
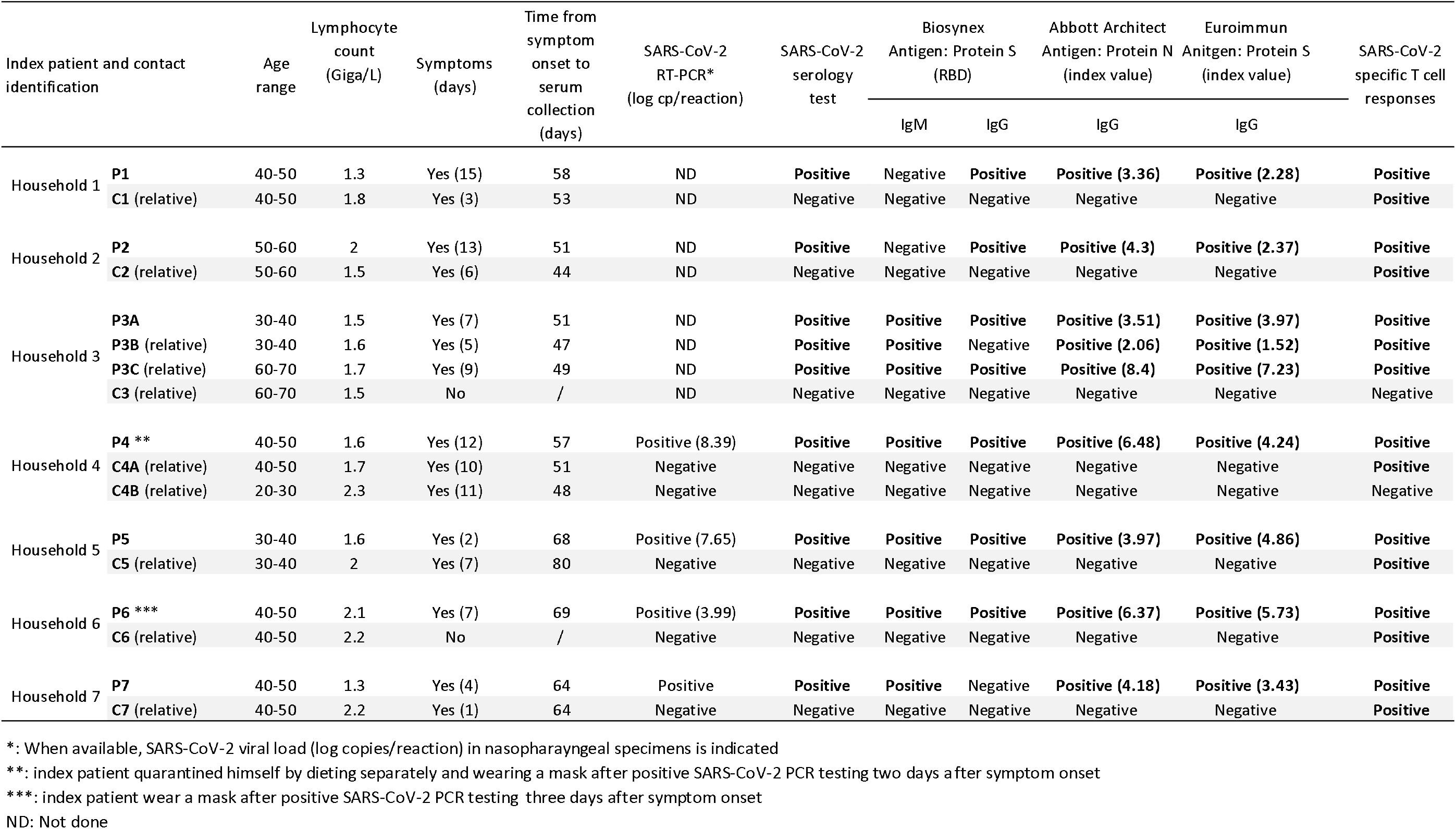
Household clinical and virological characteristics.

Serological status for SARS-CoV-2 was confirmed for index patients and contacts by using three different serological assays (Table 1). To determine whether SARS-CoV-2 specific T cells were induced in index patients and contacts, fresh PBMC samples were stimulated with structural and accessory SARS-CoV-2 proteins followed by IFNγ ELISPOT analysis. All index patients showed SARS-CoV-2-specific IFNγ responses against at least four SARS-CoV-2 antigens with a maximum of eight antigens. They all recognized the structural proteins S1, S2, N and M, and six of them recognized at least one accessory protein (3A and/or 7A, 8, 9B), suggesting that they had developed SARS-CoV-2-specific T cell responses (Fig.1A and 2). Importantly, blood samples were collected from 47 to 69 days post symptom onset, which suggests that antiviral T cells are maintained in patients recovered from a mild COVID-19 up to 69 days post symptoms.

**Figure 1.**
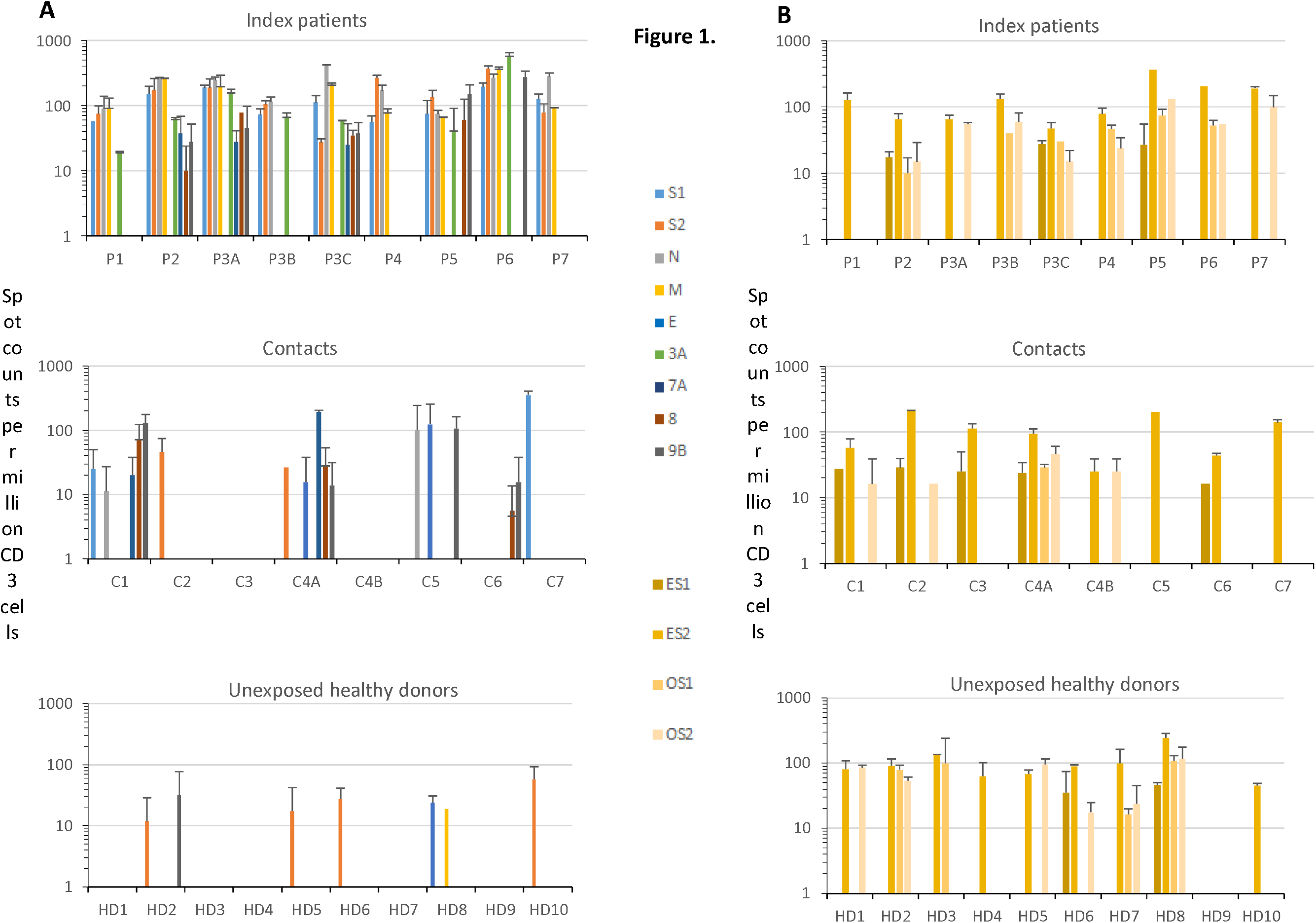
Frequency of IFN-⍰ spot forming cells against SARS-CoV-2 and two common human coronaviruses antigens in index patients, contacts and unexposed healthy donors. Means of spot counts of IFN-⍰ T cell response per million of CD3 are indicated. The experiments were performed in duplicates. T-cell secretion of IFN-⍰ was determined against (A) the SARS-CoV-2 structural proteins S1, S2, N, M and the accessory proteins 3A, 7A, 8 and 9B and against (B) the structural proteins of the human coronaviruses HCoV-229E (ES1, ES2) and HCoV-OC43 (OS1, OS2). Each color correspond to one antigen. P, index patient; C, contact; HD, unexposed healthy donor.

**Figure 2.**
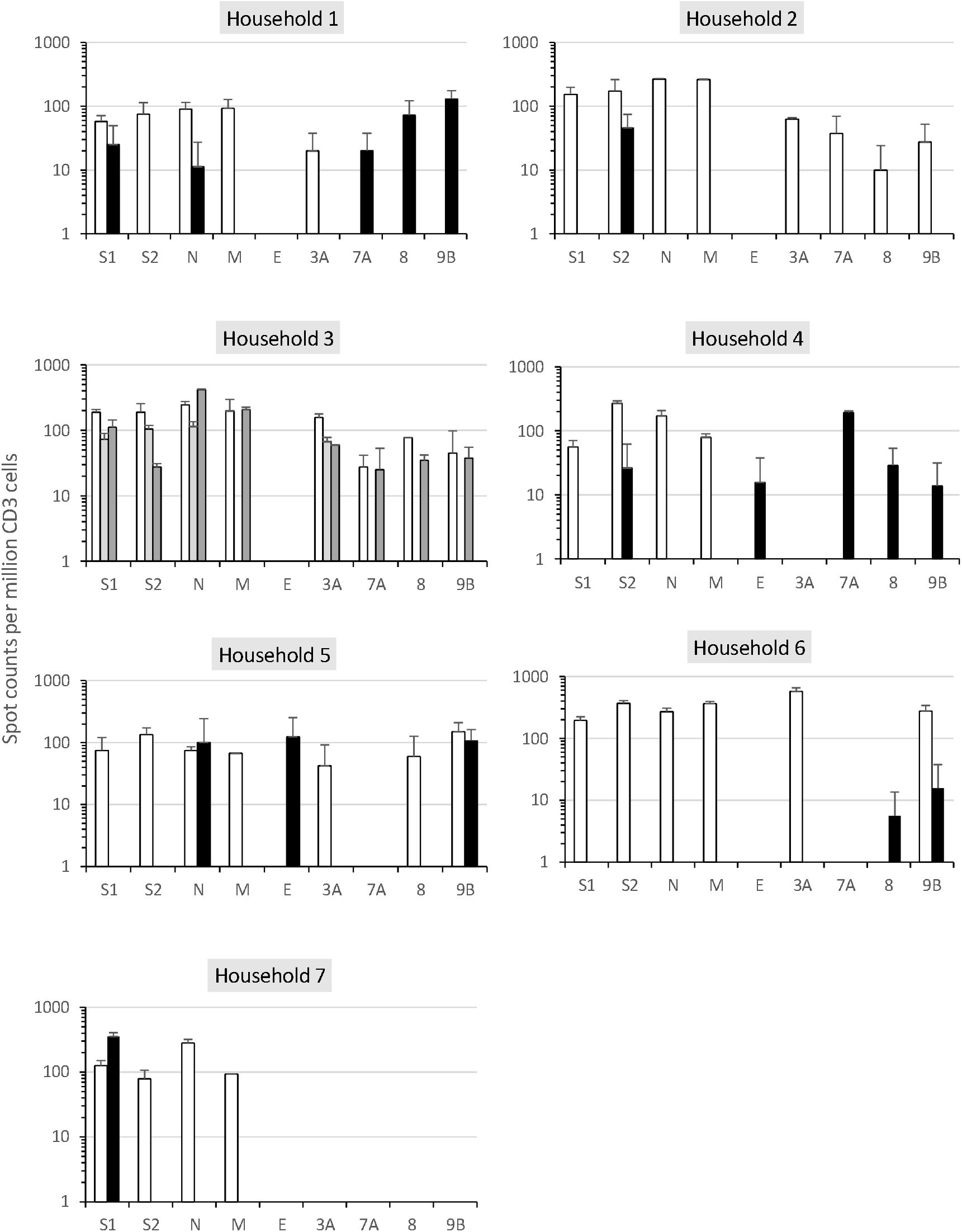
SARS-CoV-2-specific T cell response pattern in households. Means of spot counts of IFN-⍰ T cell response per million of CD3 against SARS-CoV-2 antigens in each household are shown. The X axis represent the SARS-CoV-2 antigens. The experiments were performed in duplicates. White and grey bars correspond to T cell responses detected in index patients and black bars correspond to those detected in contacts. P, index patient, C, contact.

Six of eight contacts demonstrated SARS-CoV-2 –specific IFNγ responses against at least one SARS-CoV-2 antigen (Fig.1A and 2). The contacts C1 and C4A exhibited a T-cell reactivity against five SARS-CoV-2 antigens including two structural proteins and three accessory proteins; the contact C5 exhibited a T-cell reactivity against the two structural proteins N and E and the accessory protein 9B; the contacts C2, C6 and C7 exhibited a T-cell reactivity against S2, 8/9B and S1, respectively. Of note, the frequencies of IFNγ-secreting T cells in contacts C1, C2, C4A, C5 and C7 were similar to those displayed by the index patients and much higher than those detected in unexposed healthy donors (HD) except for the antigen S2 for which HD10 displayed a similar frequency as for index patients and contacts (Fig. 1A and 2). This suggests that the contacts had developed SARS-CoV-2-specific T cell responses.

We tested index patients, contacts and unexposed healthy donors for the spike protein (pools S1 and S2) of HCoV-229E and HCoV-OC43. They all but one unexposed healthy donor (HD9) showed a high frequency of IFNγ-producing T cells directed against HCoV 229E and/or HCoV OC43 (Figure 1B).

## Discussion

In this study, for the first time, we demonstrate that intrafamilial contacts can display a SARS-CoV-2-specific T cell response in the absence of seroconversion. This T cell response provides evidence that i) transient and/or anatomically contained SARS-CoV-2 infection must have occurred and ii) T cell responses may be more sensitive indicators of SARS-Co-V-2 exposure than antibodies.

Index patients and contacts showed different T cell immunodominance patterns. The structural proteins S1, S2, M and N were clearly co-dominant and recognized by 100% of index patients. Significant T-cell responses were also directed against accessory proteins. A similar pattern was reported for SARS-CoV-2-positive patients by others [12, 13]. For the first time, our data demonstrate that the mild forms of COVID-19 induce a significant T cell response detectable at least 69 days after symptom onset.

Five of the six contacts who developed SARS-CoV-2-specific T cell responses were symptomatic. Although their reactivity pattern was mainly restricted to one structural protein and/or nonstructural proteins, the SARS-COV-2 positive T cell responses observed in contacts showed similar frequencies of SARS-CoV-2 IFNγ-producing T cells as compared to the index patients, which suggests that they had developed SARS-CoV-2-specific T cell responses. Recently, by using a T cell receptor-dependent Activation Induced Marker assay, Grifoni et al reported a CD4+ T cell response in 40-60% of unexposed individuals especially against the S protein of SARS-CoV-2. They suggested that this T cell response results from cross-reactivity with other coronaviruses [12]. While the amplitude of the T cell response (including CD4+ and CD8+ T cells) specific to common coronaviruses, including the highly prevalent HCoV-229E and the less prevalent HCoV-OC43 in France [14], were similar in index patients, contacts and unexposed healthy donors, frequencies of SARS-CoV-2-specific IFNγ^+^ T cells were much higher in index patients and contacts. These data indicate that the SARS-CoV-2-specific T cell response detected in the contacts is specific and not only a result of a cross-reactivity with common coronaviruses.

The observation of a detectable viral-specific T cell response without seroconversion is reminiscent of what was observed for occupational or household exposure to hepatitis C where subclinical transmission of HCV resulted in priming of a T-cell HCV-specific response in the absence of antibodies [6, 7]. There are multiple explanations for the development of viral-specific T cells without an antibody response. These include exposition to low doses of the virus with brief and transient viral replication, a downstream event of protective innate immune responses or abortive replication of defective viral genomes. Nevertheless, it is not clear whether these T cell responses without antibodies provide protection against a reinfection.

Although our study has a limited sample size, the results reveal that individuals exposed to SARS-CoV-2 may develop virus-specific T cell responses without antibodies. This newly discovered aspect of the immune response against SARS-CoV-2 will significantly contribute to the understanding of the natural history of COVID-19. Furthermore, our data indicate that epidemiological data relying solely on the detection of SARS-CoV-2 antibodies may lead to a substantial underestimation of prior exposure to the virus. Our data may also have implications for vaccine development and tracking the future evolution of the SARS-CoV-2 pandemic.

## Data Availability

data can be provided by the corresponding author

## Abbreviations

COVID-19: Corona Virus Disease 2019
SARS-CoV-2: severe acute respiratory syndrome coronavirus 2
HCoV: human coronaviruses
RT-PCR: reverse transcriptase-polymerase chain reaction
RBD: Receptor Binding Domain
IFN: interferon
PBMCs: Peripheral blood Mononuclear Cells
HD: Healthy Donors

## Acknowledgements

We thank all the participants to this study. This study was supported by the Strasbourg University Hospital [COVID-HUS study-HUS N°7760].

## References

1. Li Q, Guan X, Wu P, et al. Early Transmission Dynamics in Wuhan, China, of Novel Coronavirus-Infected Pneumonia. The New England journal of medicine 2020; 382:1199–207.

2. Studdert DM, Hall MA. Disease Control, Civil Liberties, and Mass Testing - Calibrating Restrictions during the Covid-19 Pandemic. The New England journal of medicine 2020.

3. Lescure FX, Bouadma L, Nguyen D, et al. Clinical and virological data of the first cases of COVID-19 in Europe: a case series. The Lancet Infectious diseases 2020; 20:697–706.

4. Kissler SM, Tedijanto C, Goldstein E, Grad YH, Lipsitch M. Projecting the transmission dynamics of SARS-CoV-2 through the postpandemic period. Science 2020; 368:860–8.

5. Hall MA, Studdert DM. Privileges and Immunity Certification During the COVID-19 Pandemic. Jama 2020.

6. Freeman AJ, Ffrench RA, Post JJ, et al. Prevalence of production of virus-specific interferon-gamma among seronegative hepatitis C-resistant subjects reporting injection drug use. J Infect Dis 2004; 190:1093–7.

7. Heller T, Werner JM, Rahman F, et al. Occupational exposure to hepatitis C virus: early T-cell responses in the absence of seroconversion in a longitudinal cohort study. J Infect Dis 2013; 208:1020–5.

8. Mizukoshi E, Eisenbach C, Edlin BR, et al. Hepatitis C virus (HCV)-specific immune responses of long-term injection drug users frequently exposed to HCV. J Infect Dis 2008; 198:203–12.

9. Bryan A, Pepper G, Wener MH, et al. Performance Characteristics of the Abbott Architect SARS-CoV-2 IgG Assay and Seroprevalence in Boise, Idaho. J Clin Microbiol 2020.

10. Beavis KG, Matushek SM, Abeleda APF, et al. Evaluation of the EUROIMMUN Anti-SARS-CoV-2 ELISA Assay for detection of IgA and IgG antibodies. Journal of clinical virology: the official publication of the Pan American Society for Clinical Virology 2020; 129:104468.

11. Fafi-Kremer S, Bruel T, Madec Y, et al. Serologic responses to SARS-CoV-2 infection among hospital staff with mild disease in eastern France. medRxiv 2020:2020.05.19.20101832.

12. Grifoni A, Weiskopf D, Ramirez SI, et al. Targets of T Cell Responses to SARS-CoV-2 Coronavirus in Humans with COVID-19 Disease and Unexposed Individuals. Cell 2020.

13. Ni L, Ye F, Cheng ML, et al. Detection of SARS-CoV-2-Specific Humoral and Cellular Immunity in COVID-19 Convalescent Individuals. Immunity 2020.

14. Lepiller Q, Barth H, Lefebvre F, et al. High incidence but low burden of coronaviruses and preferential associations between respiratory viruses. J Clin Microbiol 2013; 51:3039–46.

